# General Framework for Evaluating Outbreak Prediction, Detection, and Annotation Algorithms

**DOI:** 10.1101/2022.03.16.22272469

**Authors:** Auss Abbood, Stéphane Ghozzi

## Abstract

The COVID-19 pandemic has highlighted and accelerated the use of algorithmic-decision support for public health. The latter’s potential impact and risk of bias and harm urgently call for scrutiny and evaluation standards. One example is the early detection of local infectious disease outbreaks. Whereas many statistical models have been proposed and disparate systems are routinely used, each tailored to specific data streams and use, no systematic evaluation strategy of their performance in a real-world context exists.

One difficulty in evaluating outbreak prediction, detection, or annotation lies in the scales of different approaches: How to compare slow but fine-grained genetic clustering of individual samples with rapid but coarse anomaly detection based on aggregated syndromic reports? Or alarms generated for different, overlapping geographical regions or demographics?

We propose a general, data-driven, user-centric framework for evaluating heterogeneous outbreak algorithms. Discrete outbreak labels and case counts are defined on a custom data grid, associated target probabilities are then computed and compared with algorithm output. The latter is defined as discrete “signals” are generated for a number of grid cells (the finest available in the benchmarking data set) with different weights and prior outbreak information from which then estimated outbreak label probabilities are derived. The prediction performance is quantified through a series of metrics, including confusion matrix, regression scores, and mutual information. The dimensions of the data grid can be weighted by the user to reflect epidemiological criteria.

## 1 Introduction

In epidemiology of infectious diseases, one typically distinguishes between endemic and epidemic infection dynamics [1]. In the first, cases are widely spread among a population and broad control measures are recommended. Epidemics, i.e., dynamical spread of diseases, on the contrary, can be best controlled with rapid and targeted measures. Thus, understanding the epidemic potential of a disease and rapidly identifying local epidemics can be beneficial public health tools. This has been vividly illustrated by the track for COVID-19 clusters.

Here, we propose a general approach to evaluate such tools. We use the term “outbreak” to designate a set of infection cases epidemiologically linked one to the other. This can be because widely distributed food has been contaminated by salmonella, dengue-carrying mosquitoes are spreading, one SARS-CoV-2 infected persons meets others in a closed space, or, in the classical example, through one source of contaminated water causing cholera. An outbreak is defined by experts, e.g., after investigation or through predefined rules.

Many algorithms have been proposed for the tasks of identifying outbreaks in the past (annotation), currently happening (detection), or expected to happen (prediction) [2–6]. Some of them have been routinely used by public health agencies worldwide for years [7–9]. However, little attention has been paid to their systematic evaluation and such work was mostly limited to simple simulations, very specific settings and types of algorithms [9–12]. Beyond algorithm evaluation, this framework could serve as basis for hyperparameter optimization [13], as well as data visualization and semisupervised learning [14].

In this paper, we outline a way to define a benchmarking data set based on reported infection cases and a general heuristic, inspired by standard machine-learning practices and based on the needs of public health agents.

Different types of algorithms can be evaluated and compared through a series of standard scores, which can be weighted to reflect domain expert priorities. Typically those would apply time-series analysis to aggregated case numbers, one challenge being that cases can be aggregated in many different ways. Moreover, outbreaks can overlap. Thus, different scales have to be considered at the same time.

## 2 Benchmarking data set

### 2.1 Cases and outbreaks

We consider individual infection cases reported to a public health agency. Each case is assigned *data labels* that either identify the outbreaks to which it belongs (in the following illustrated as numbers, 1, 2, 3, …, *N*) or that it doesn’t belong to any outbreak (endemic or sporadic, and identified in the following with ⊗). Exogenous data used for training algorithms or for predictions such as weather, mobility patterns, or other determinants, are not part of the benchmarking data set.

The set of corresponding data labels is denoted by ***d*** = {*d*_1_, *d*_2_, …}. One case can have many labels if there is uncertainty on its status. More precisely, all available labels are assigned to each case with different probabilities, i.e., let p_*c*_(*d*) be the probability that a case *c* has label *d* with the constraint that ∀*c*, Σ_*d*_ p_*c*_(*d*) = 1. Uncertainty can be caused by the labelling method, e.g., PCR tests, or by the reporting system, e.g., the extent to which the case definitions are met.

### 2.2 Data grid and cells

Each reported infection case has a number of properties relevant to the public health agency. We define a *data grid* comprised of *data cells*, points in the space defined by the highest resolution of available labeled case properties. The coordinates of one given cell is denoted by ***x*** = {*x*_1_, *x*_2_, …, *x*_*D*_}. Properties can be quantitative (age, infection date, body temperature), in which case they need to be discretized, or categorical (sex, symptoms). For example, if cases have the features date, place, sex measured as calendar week, county, female/inter-sexual/male, one cell might have the coordinate {2020-CW2, Berlin, female}. The dimensions need not be restricted to epidemiological, demographic or genetic data but, given an one-health approach, could also include data on animal reservoirs or environmental samples.

Empty cells are assigned a single dummy “non-case” denoted by ∅. Thus, considering both outbreak, endemic and non-cases labels, there are overall *N* + 2 data labels.

In the following, data cells are considered as the units of evaluation, see Fig. 1.

**Figure 1:**
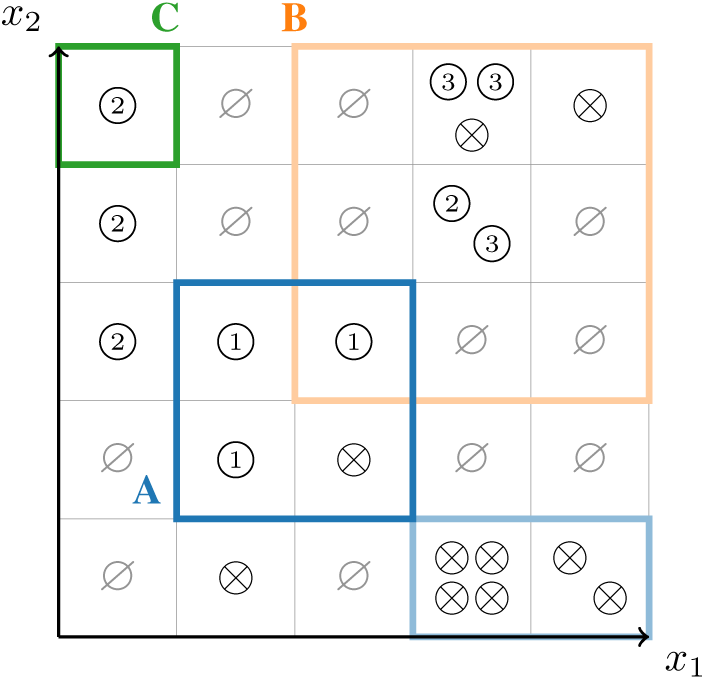
Data cells of a two-dimensional data grid, data labels, and signals. The circles represent cases and the grid the boundaries of data cells. The data classes are {1, 2, 3, ⊗, ∅ }. The signal classes are {A, B, C, ⊗, ∅ }. For illustration purposes “interesting” signals are shown as frames around cells with color saturation corresponding to *w*(*s*, ***x***), and frames not shown when this probability is 0. The signals ⊗ and ∅ are not shown.

### 2.3 Data cell labels and outbreak probabilities

Based on case labels, data cells themselves are assigned labels. The expected number of cases belonging to class *d* in a given cell ***x*** is the sum of the corresponding probabilities: 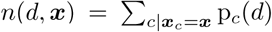. We ignore the possibility of uncertainty on the coordinates of a case, so that any case *c* can be attributed unambiguously to a given cell ***x***_*c*_.

p(*d*|***x***), the probability for cell ***x*** to have label *d*, is defined as the probability of drawing a case with label *d* from cell ***x***: p(*d*|***x***) = *n*(*d*, ***x***)/Σ_*d*′ *′*_*n*(*d*′, ***x***).

## 3 Signals and algorithm output

We define a *signal* as the output of an algorithm indicating a possible outbreak. Signals are identified by *signal labels* denoted by ***s*** = {*s*_1_, *s*_2_, …, *s*_*M*_}, here by letters A, B, C, … for the outbreak signals, together with endemic signals ⊗ and empty signals ∅. One algorithm might generate many signals, e.g., because it considers different aggregation scales or because it takes into accounts that many outbreaks have been already observed.

We assume that signals are generated with a certain measure of significance or importance, which might vary across cells, denoted by *w*(*s*, ***x***). It can be for example a probability of belonging to an outbreak, a p-value of rejecting the null hypothesis “endemic”, or simply binary (0 or 1). Some algorithm might operate at a coarser level than available in the data set, e.g., it could aggregate cases weekly to avoid the weekend effect when in fact cases are reported daily. In this case, we simply assume *w*(*s*, ***x***) to be constant over all the corresponding cells. Fig. 1 illustrates signal weights on the data grid.

For the sake of generality, we expect *w* to be a real number between 0 and 1, and to always be defined for endemic and empty signals. In practice, this means ad-hoc transformations, such as scaling, might need to be applied to algorithm output. Note that this means that for each cell at least one *w* will be non-zero.

We propose a heuristic to define a “probability” p(*s*|***x***) for a data cell *x* to be attributed to a signal *s*. We use the word probability without any pretense of sound statistical meaning but solely for illustrating the idea of the approach. This quantity is simply taken to be the weight of a signal relative to the sum of all weights: p(*s*|***x***) = *w*(*s*, ***x***)/Σ_*s*′_ *w*(*s*′, ***x***).

Lastly, such signals might incorporate information on previously known outbreaks. Moreover, we assume “endemic” and “non-case” signals are produced by all algorithms and that these labels have the same meaning as their counterparts in the data, leading to overall *M* + 2 signal labels. This a-priori relationship between signal and data label, i.e., the information on data labels considered by the algorithm, is defined by the latter and denoted here by p(*d*|*s*). E.g., an algorithm that doesn’t use data labels for training would have the following priors: p(*d* = ⊗|*s* =) = p(*d* = ∅|*s* = ∅) = 1; p(*d* ≠ ⊗|*s* =) = p(*d* ≠ ∅|*s* = ∅) = p(*d* = ⊗|*s* ≠ ⊗) = p(*d* ≠ ∅|*s* = ∅) = 0; and p(*d*|*s*) = 1*/N* for all other combinations of data and signal labels. Here again, we use the notation of probabilities but stress the heuristic nature of our approach.

Combining signal probabilities and prior information on outbreaks, inspired by Bayes’s formula, we define an estimator for the probability of data cell ***x*** having label *d*: 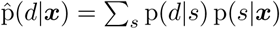.

## 4 Evaluation

### 4.1 Standard machine-learning scores

The overlap of signal and data labels can be investigated in a number of ways. Cross-entropy can be computed as 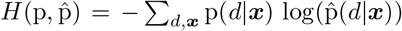 (in practice, here one might have to address logs of zero [15], e.g., by adding a small constant to 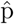). One can also compute joint probabilities: p(*d, s*) = p(*d*|*s*) p(*s*) = Σ_***x***_ p(*d*|*s*) p(*s*|***x***)/|{***x***}|, or, equivalently, p(*d, s*) = p(*s*|*d*) p(*d*) = Σ_***x***_ p(*s*|*d*) p(*d*|***x***)/|{***x***}|, where ***x*** is the number of data cells. These can help interpretation through visualization as well as derived quantities. For example, one can define a mutual information between signals and data: *I* = Σ_*d,s*_ p(*d, s*) log(p(*d, s*)/p(*d*)p(*s*)).

Estimating the agreement of signal and data labels can be performed as a classification or as a regression task. By setting thresholds on p(*d*|***x***) and 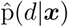, one can construct a confusion matrix and derive recall, precision, F-score, etc., as well as binary cross-entropy, AUC or Brier score. Multiclass and multilabel scores can be defined after one-hot encoding of p(*d*|***x***) (e.g., via most probable class) and replication of each cell (*N* times and then applying the threshold), respectively [16]. Seen as regression, one can compute distances between the two probabilities, e.g., mean absolute error or quantiles.

### 4.2 Epidemiological motivation

Some performance properties might be of particular importance for epidemiology or epidemic preparedness and response. For example, timeliness is, in practice, an important factor in outbreak detection: The sooner one is aware of an epidemic, the better the response can be. This can be considered by reducing the data grid to one time dimension (projecting all cases on that dimension) and computing a function that vanishes before the first outbreak case, then equals one and, with a given time scale, vanishes with time.

More generally, all scores considered above include sums over all data cells. These sums can be weighted to reflect the importance of certain aspects. E.g., we can define space-time weights [3, 17–20] as follows: For each outbreak case, a multi-dimensional function that monotonically vanishes with increasing spatial and time distance, together with a mask that is 0 for all times before the first case, and 1 afterwards. This defines, for each data label, case, and cell a weight between 0 and 1. Normalizing these over all cases leads to label and cell specific weights.

Other weights can be defined to account for vulnerable populations (by weighting along a given dimension) or spreading properties (network distance from the index case on the chains of infection). Risk for discrimination can be further measured or accounted for through Rawlsian maximin approach [21], as well as other post-processing approaches [22].

## 5 Conclusion

We have shown how to compute evaluation scores for the problem of infection outbreak prediction, detection, and annotation. This is done on a benchmarking data set that is provided by an interested party, e.g., a public health agency. This thus ensures automatically that the benchmarks are relevant for the specific context in which algorithms are expected to be used. Furthermore, outbreaks can be defined in any epidemiologically sound and operational way by public health agents. We hope this work can form the basis for a public-health standard, where algorithms to support decision making are scrutinized and evaluated in relevant contexts [23].

Future work will include applying this framework to simulated and real data. One special use case that will require specific attention is the combination of classical epidemiology and phylogenetics. Uncertainties in case coordinates, labelling, and even case numbers will need to be addressed. This would help establish proper probabilistic foundations, potentially important in the context of Bayesian optimization.

The simple scoring approach presented here is a small part of a proper ethical and social evaluation. All aspects of algorithm usage should be considered [24]. Representativity and quality of data sets, choice of training process and loss functions, and who is able or willing to use the algorithms and what for, are all relevant to the desirability of algorithm-supported public-health decision taking.

## Data Availability

No data is part of this work.

## Code availibility

The framework described here is implemented in the open-source Python package epi-quark: https://github.com/aauss/epi-quark

## Broader impact

We expect our work to help mitigate the risk of a “one-size-fits-all” practice of evaluating algorithms in the specific context (country, reporting system) in which they were developed. However, the focus on quantitative scores might reinforce unwarranted trust in algorithmic analyses, too often seen as objective, neutral, and universal.

## Acknowledgements

We thank Rachel Lowe for helpful discussions.

